# Sequential Word Properties in Verbal Fluency: Detecting High-Proficiency Cognitive Impairment

**DOI:** 10.64898/2026.07.06.26357360

**Authors:** Ya-Ning Chang, Yi-Hsuan Wang, Chia-Ju Chou, Yi-Chien Liu, Matthew A. Lambon Ralph

## Abstract

Verbal fluency (VF) tasks are widely used to differentiate patients with cognitive impairment from healthy controls, but total word count produced during these tasks becomes unreliable when patients and controls exhibit comparable proficiency. This study examined, in detail, whether item-level and sequential properties of words produced during a VF task could reliably differentiate high-proficiency patients indistinguishable from controls by word count alone. Seventy-seven native Mandarin Chinese speakers (38 controls and 39 patients with mild cognitive impairment or mild dementia) completed a semantic VF task. Participants were subdivided by proficiency into four groups: high-proficiency controls (HC), low-proficiency controls (LC), high-proficiency patients (HP), and low-proficiency patients (LP). The LC and HP subgroups were matched on semantic fluency scores and thus provided a key focus for the investigation. We examined item-level properties (word frequency, contextual diversity, semantic diversity, surprisal) and sequential properties (positional frequency variation) of the words produced. Significant group differences emerged across item-level psycholinguistic properties, though these were primarily driven by the LP group, with no reliable differentiation between LC and HP. Crucially, positional frequency variation distinguished LC from HP. LC participants began their lists with high-frequency words followed by a systematic decline, whereas HP patients produced words within a consistently narrow frequency band throughout. These findings indicate that item-level psycholinguistic properties alone are insufficient to differentiate HP from LC, whereas sequential word frequency variation provides a potential index of cognitive impairment, reflecting underlying differences in semantic retrieval and memory organisation. Future work with larger samples is needed to validate generalisability.

## 1. Introduction

Verbal fluency (VF) tasks have been widely used since the 1960s to detect neurodegenerative diseases such as Alzheimer’s disease (AD) and its prodromal stage, mild cognitive impairment (MCI) (Benton, 1968; Binder & Desai, 2011; Patterson et al., 2007; Vonk et al., 2019). VF has been used to evaluate semantic memory and executive function (Henry & Crawford, 2004), and subsequent research has increasingly identified its sensitivity to early AD (McDonnell et al., 2020; Mueller et al., 2015).

Traditional VF assessments often focus on total word count. Given their straightforward administration and effectiveness, they are now routinely used as a global screen for neurodegenerative diseases. However, this approach has several notable limitations. For example, in a large-scale study of six neurodegenerative diseases, Henderson et al. (2023) evaluated the clinical utility of VF tests using a range of psycholinguistic analyses (e.g., word properties and production order). They demonstrated that despite providing good differentiation of all patient groups against controls, VF has limited utility in differentiating between various patient groups – even those that are clearly clinically different such as AD versus behavioural variant of frontotemporal dementia (bvFTD). The only partially individuated group were those with semantic dementia (SD)/semantic variant of primary progressive aphasia (svPPA). By going beyond simple word count alone, SD/svPPA patients were shown to have a disproportionate impairment in category over letter fluency and tended to produce higher-frequency words. This contrastive pattern of results (good patient vs. control differentiation but poor intra-patient resolution) probably reflects the fact that VF relies on a wide array of cognitive functions (Henderson et al., 2023; Lambon Ralph et al., 2003), including attention, memory (Greenberg et al., 2009), executive function (Aita et al., 2019) and speed of lexical retrieval (Elgamal et al., 2011), any of which, if impaired, compromises performance regardless of the specific disease.

A further limitation arises when individuals produce similar numbers of words, as simple counting makes it difficult to detect subtle impairments. Therefore, beyond traditional quantitative measures, researchers have sought qualitative analyses to understand item-level variation, including the difficulty of produced words (Murphy & Castel, 2021), age of word acquisition (Forbes-McKay et al., 2005), clustering and switching patterns (Troyer et al., 1997; Weakley & Schmitter-Edgecombe, 2014), the word properties of the first five words produced (Forbes-McKay et al., 2005; Wakefield et al., 2018) and network analysis (Bertola et al., 2014; Lerner et al., 2009; Santos et al., 2017). These approaches capture the information inherent in individual words. However, most existing studies has focused on Western languages, particularly English. As word properties can vary considerably across languages, which properties are most diagnostic for discriminating high-proficiency patients from controls remain unclear for some understudied yet commonly spoken languages such as Mandarin Chinese.

The English and Mandarin Chinese language systems are different in orthographic, phonological, and semantic processing, as reflected in different degrees of orthographic-to-semantic mappings and the extent of semantic mediation in lexical processing (e.g., Chang & Lee, 2020; Perfetti et al., 2005; Perfetti & Tan, 1998; Tan & Perfetti, 1998). Although several psycholinguistic measures are conceptually comparable across the two languages, such as word frequency, semantic diversity, and contextual diversity, they are not identical in their specific instantiation due to structural differences between the two language systems (Cai & Brysbaert, 2010; Liu et al., 2007; Chang et al., 2016). A key phonological difference is that Mandarin Chinese is a tonal language, in which pitch is used to signal differences in word meaning, whereas English is non-tonal (e.g., Cheng, 2011; Crinion et al., 2009; Gandour et al., 2000; Howie, 1976). These language-specific factors may influence VF performance (Luo et al., 2010; Rosselli et al., 2002). In addition, concepts and their associations (which contribute to category fluency production) vary between cultures. For example, relevant for the animal fluency task explored in this study and used routinely in clinical settings, not only do exemplars vary in familiarity across world locations but also cultures have additional associations between them beyond typical animate hierarchies (e.g., the zodiac-association between diverse animals).

Consequently, it is important to investigate the influence item-level properties on differentiating Mandarin Chinese-speaking patients with cognitive impairment from healthy controls particularly in cases where patients and healthy individuals exhibit comparable word production efficiency. Therefore, in this study, we investigated whether item-level and sequential properties of words produced during a VF task can reliably detect cognitive impaired patients, even those with relatively high verbal fluency (overlapping with the lower end of normative scores). Specifically, we investigated whether healthy controls and patients differ in item-level and sequential properties of the words they produced; and, if so, whether the identified properties could be used to train a classifier to differentiate between groups, and critically, whether it could generalise to out-of-sample data.

## 2. Method

### 2.1. Participants

Seventy-seven Mandarin Chinese speaking participants were recruited from the memory clinic of Cardinal Tien Hospital in Taipei, Taiwan, between September 2019 and August 2023. Inclusion/exclusion criteria included: aged between 60 and 90 years, at least six years of education, and no history of neurological or psychiatric conditions (e.g., stroke, head injury, or brain tumour). Participants were additionally required to have been followed at the memory clinic for at least one year and to have received a diagnosis of cognitive impairment within two years of study enrolment.

Patients were primarily categorised into two groups based on Clinical Dementia Rating (CDR), Clinical Dementia Rating-Sum of Boxes (CDR-SB), and Mini-Mental State Examination (MMSE) scores, with the final classification determined by the senior neurologist (Y.C.L.) through the integration of cognitive scores and clinical observations. The patient group comprised 39 individuals with MCI and mild dementia, and the control group comprised 38 healthy individuals.

For each participant group, we further divided participants into two subgroups based on word generation proficiency (i.e., number of unique words produced), resulting in four groups: 21 high-proficiency controls (HC), 17 low-proficiency controls (LC), 20 high-proficiency patients (HP) and 19 low-proficiency patients (LP). By design, the number of unique words produced did not differ significantly between the LC and HP groups (*p* > 0.05) and thus provided a key challenge for the investigations – which fluency measures beyond simple word count, if any, can differentiate between them. Table 1 presents detailed demographic information for all four groups.

**Table 1.**
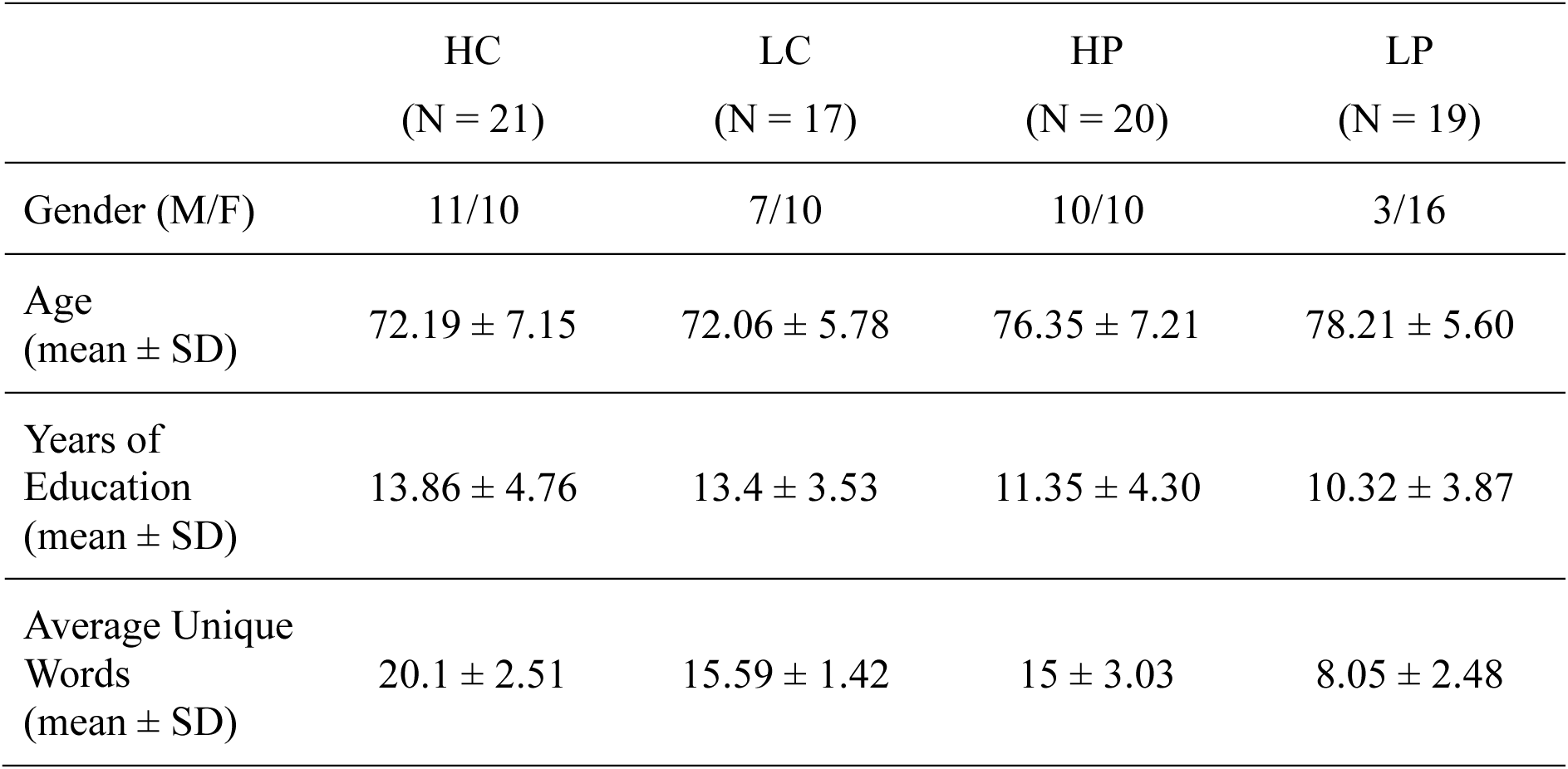
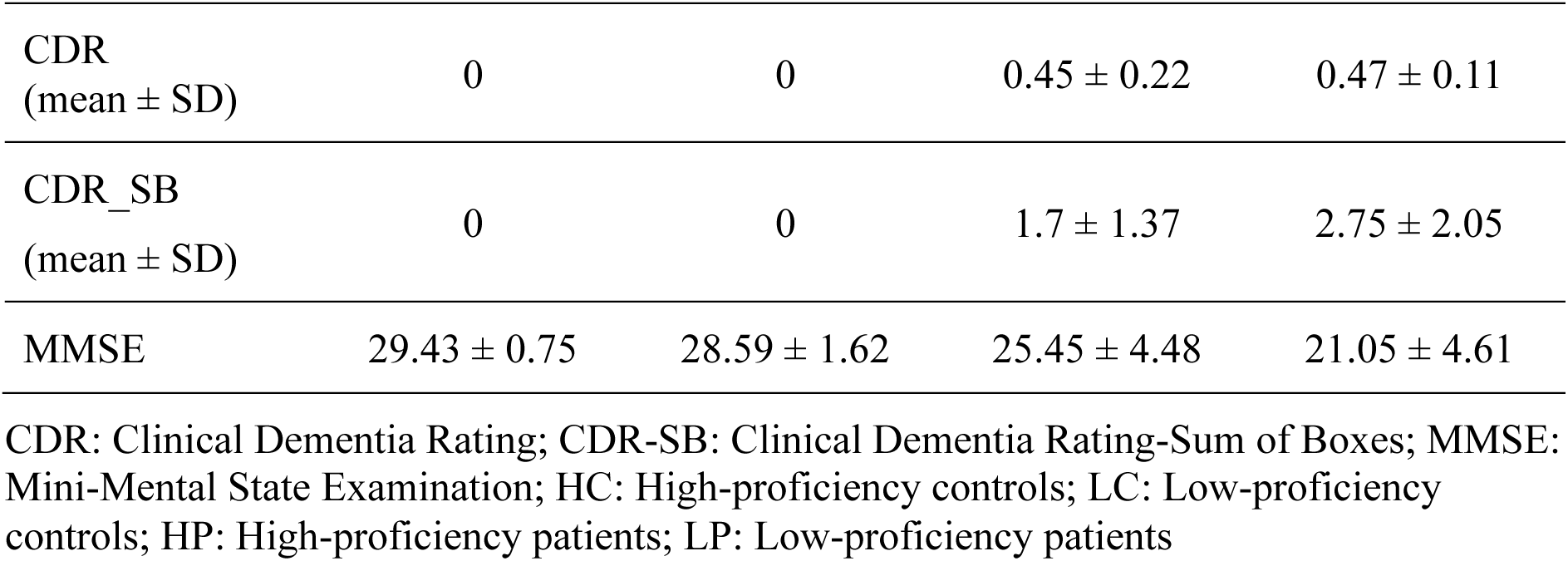
The descriptive information for the participants.

|  | HC<br>(N = 21) | LC<br>(N = 17) | HP<br>(N = 20) | LP<br>(N = 19) |
| --- | --- | --- | --- | --- |
| Gender (M/F) | 11/10 | 7/10 | 10/10 | 3/16 |
| Age<br>(mean ± SD) | 72.19 ± 7.15 | 72.06 ± 5.78 | 76.35 ± 7.21 | 78.21 ± 5.60 |
| Years of<br>Education<br>(mean ± SD) | 13.86 ± 4.76 | 13.4 ± 3.53 | 11.35 ± 4.30 | 10.32 ± 3.87 |
| Average Unique<br>Words<br>(mean ± SD) | 20.1 ± 2.51 | 15.59 ± 1.42 | 15 ± 3.03 | 8.05 ± 2.48 |
| CDR<br>(mean ± SD) | 0 | 0 | 0.45 ± 0.22 | 0.47 ± 0.11 |
| CDR_SB<br>(mean ± SD) | 0 | 0 | 1.7 ± 1.37 | 2.75 ± 2.05 |
| MMSE | 29.43 ± 0.75 | 28.59 ± 1.62 | 25.45 ± 4.48 | 21.05 ± 4.61 |

CDR: Clinical Dementia Rating; CDR-SB: Clinical Dementia Rating-Sum of Boxes; MMSE: Mini-Mental State Examination; HC: High-proficiency controls; LC: Low-proficiency controls; HP: High-proficiency patients; LP: Low-proficiency patients

### 2.2. Data collection and preprocessing

Verbal fluency tests were conducted at the memory clinic. Participants were asked to produce as many words as possible from the animal category within one minute, without repetition. Audio recordings were processed using a semi-automated pipeline. First, the recordings were automatically transcribed using Whisper medium (https://github.com/openai/whisper). Second, a rule-based approach was applied to extract target responses. The rules were as follows: the transcript was segmented using common examiner prompts and cue words (e.g., *begin*, *ready*, and *tell me*); non-category words that frequently appeared in the transcripts were removed, including terms such as *test*, *thing*, *mammal*, and *animal*; the cleaned text was subsequently tokenised using the Academia Sinica CKIP model (https://github.com/ckiplab/ckiptagger) to extract content nouns. To simplify the analysis, items with compound adjective-noun forms such as *澳洲梨 (Australian pear)* and *鳳梨釋迦 (pineapple custard apple)* were reduced to their noun components. Finally, two types of errors were identified and excluded: (1) words that participants whispered, self-repeated, or produced to confirm with the examiner whether a response was appropriate, as these behaviours were unrelated to the memory retrieval process; and (2) words that did not belong to the animal category (e.g., *banana*).

### 2.3. Distributional analyses

We then conducted distribution analyses to investigate psycholinguistic word properties, including word frequency, contextual diversity, semantic diversity, and surprisal. The first three measures were taken from existing lexical norms (Chang & Lee, 2018). Surprisal scores were computed using a custom script in which each participant’s word list was passed as a sequence to a pretrained GPT-2 model (Radford et al., 2019) (https://huggingface.co/docs/transformers/en/model_doc/gpt2). For each word in the sequence, the preceding words served as the context. The probability of the target word given that context was obtained from the model’s output logits via a softmax function, and surprisal was calculated as the negative log-base-2 of that probability (i.e., -log₂*P*). For multi-token words, token-level probabilities were multiplied to yield a single word-level probability before computing surprisal. The average surprisal across all words in a participant’s list was used as the summary measure.

To compare word properties across groups, each measure was divided into three bins (Low, Medium, and High) based on quantile boundaries derived from the pooled word distribution of the control participants (40th and 70th percentiles). For each participant, the proportion of words falling into each bin was computed, such that the three bin proportions sum to 1.0 for each individual. Note that the bin boundaries were anchored to the control distribution, any systematic shift in the patient groups relative to controls reflected a departure from the normative retrieval pattern.

### 2.4. Frequency trajectories and semantic clustering analyses

In addition to item-level properties, we examined sequential patterns by tracking how word frequency values changed across serial positions within the word list, relative to the first word. Word frequency was chosen because it is one of the most powerful psycholinguistic variables across different language tasks (Balota et al., 2007; Chang et al., 2016) and has been shown to be the strongest discriminator for SD/svPPA relative to other neurodegenerative diseases (Henderson et al. 2023). Since each participant produced varying numbers of words, we focused on frequency changes across the first 15 words; for participants who produced fewer than 15 words, all available words were included. This positional frequency variation measure was used to examine whether sequential frequency patterns could reliably differentiate the HP group from the LC group.

Furthermore, to investigate shifts in word categories across the word list, a semantic clustering analysis of all words produced by participants was conducted. Specifically, each word was independently categorised by the authors and verified with the assistance of large language models (LLM; Google Gemini 2.5 Flash), based on the semantic features and habitats of animals. This resulted in eight semantic clusters: Domestic Animal (e.g., cat), Farm Animal (e.g., cow), Wild Animal (e.g., leopard), Aquatic Animal (e.g., whale), Aerial Animal (e.g., swan), Invertebrate (e.g., ant), Reptile/Amphibian (e.g., lizard), and Others (e.g., dinosaur). A representative semantic vector for each cluster was generated by averaging the word embeddings of its member words, taken from a sentence-transformers model (https://huggingface.co/sentence-transformers/distiluse-base-multilingual-cased-v1). Two sequential measures were then computed: (a) the average number of between-cluster transitions across different clusters (scored as 0 for consecutive words within the same cluster and 1 for consecutive words from different clusters); and (b) the average pairwise cluster similarity between consecutive words, derived from the cluster similarity matrix.

### 2.5. Logistic regression

To investigate whether word frequency changes in word lists could account for additional variance beyond and above simple word count, logistic regression analyses were conducted. The analyses were conducted on all participants, and on only the LC and HP groups separately, with unique word, and slope information including the slope, mean and standard deviation of word frequency changes as predictors, and group label as dependent variable (0 for healthy individuals and 1 for patients).

Furthermore, to evaluate the generalisability of the models, we additionally tested the models on an out-of-sample test of 27 participants, consisting of 2 HC, 4 LC, 6 HP, and 15 LP. The descriptive information for this out-of-sample dataset is reported in Table 2.

**Table 2.** The descriptive information for this out-of-sample dataset.

|  | HC<br>(2) | LC<br>(4) | HP<br>(6) | LP<br>(15) |
| --- | --- | --- | --- | --- |
| Gender (M/F) | 2/0 | 2/2 | 1/5 | 3/12 |
| Age<br>(mean ± SD) | 68 ± 4.24 | 72.75 ± 4.99 | 69 ± 8.07 | 73.8 ± 6.37 |
| Years of<br>Education<br>(mean ± SD) | 12 ± 8.49 | 9 ± 2.45 | 11.33 ± 4.50 | 10 ± 4.69 |
| Average Unique<br>Words<br>(mean ± SD) | 25 ± 2.83 | 13.25 ± 2.63 | 16 ± 2.28 | 8.07 ± 2.12 |
| CDR<br>(mean ± SD) | 0 ± 0 | 0.12 ± 0.25 | 0.5 ± 0 | 0.5 ± 0 |
| CDR_SB<br>(mean ± SD) | 0 ± 0 | 0.5 ± 1 | 2 ± 1 | 2.87 ± 2.35 |
| MMSE<br>(mean ± SD) | 29 ± 0 | 28.25 ± 1.5 | 21.83 ± 5.04 | 20.67 ± 5.05 |

CDR: Clinical Dementia Rating; CDR-SB: Clinical Dementia Rating-Sum of Boxes; MMSE: Mini-Mental State Examination; HC: High-proficiency controls; LC: Low-proficiency controls; HP: High-proficiency patients; LP: Low-proficiency patients

## 3. Results

### 3.1. Distribution analyses results

Figure 1 presents the mean bin proportions for word frequency, contextual diversity, semantic diversity, and surprisal across the four groups (HC, LC, HP, LP). To examine whether the four groups differed in their distributional profiles across lexical properties, series of mixed ANOVAs were conducted separately for word frequency, contextual diversity, and semantic diversity. In each analysis, group (HC, LC, HP, LP) was entered as a between-subject factor, bin (Low, Medium, High) as a within-subject factor, and the normalised proportion of unique words per bin as the dependent variable. Where significant group effects or interactions between group and bin were found, post-hoc pairwise t-tests were conducted within each bin separately, with Bonferroni correction applied to adjust for multiple comparisons.

**Figure 1.**
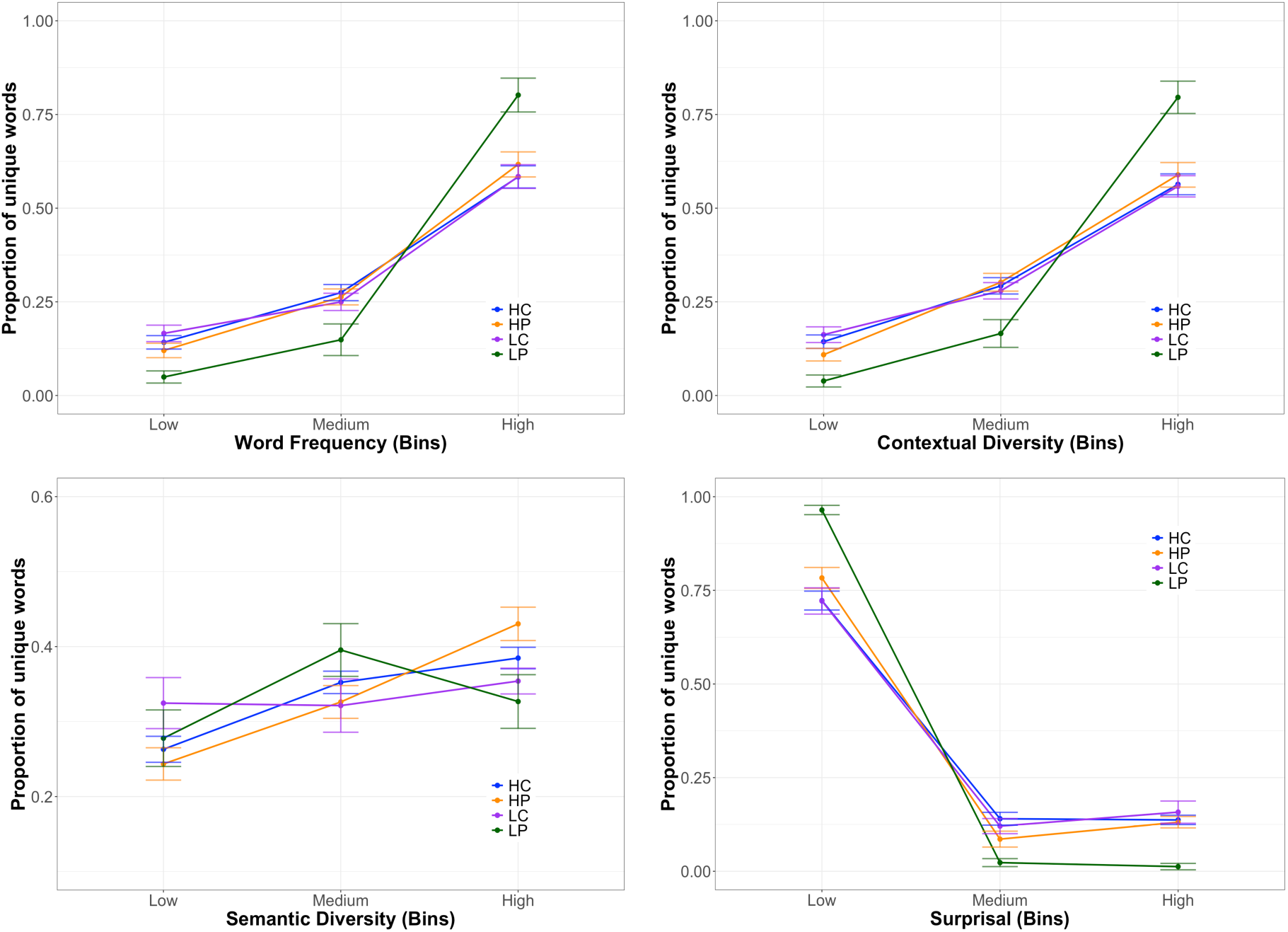
The distribution analyses of word frequency, contextual diversity, semantic diversity and surprisal for the control and patient groups. HC: high-proficiency control; LC: low-proficiency control; HP: high-proficiency patient; LP: low-proficiency patient.

For word frequency, the ANOVA results revealed a significant main effect of bin, F(2, 146) = 254.22, *p* < .001, and a significant interaction between bin and group, F(6, 146) = 6.91, *p* < .001, while the main effect of group was not significant. Post-hoc pairwise comparisons within each bin indicated that the LP group differed significantly from the other three groups, particularly in the High bin, where LP showed a notably higher proportion of words compared to HC, LC, and HP (all *ps* < 0.05). A similar but enhanced pattern was observed for contextual diversity, with a significant main effect of bin, F(2, 146) = 255.62, *p* < .001, and a significant interaction between bin and group, F(6, 146) = 9.04, *p* < .001, but no significant main effect of group. Post-hoc pairwise comparisons again showed that the LP group differed significantly from the other three groups across all three bins (all *ps* < 0.05). For semantic diversity, only the main effect of bin was significant, F(2, 146) = 9.57, *p* < .001; neither the main effect of group nor the interaction between bin and group was significant. For surprisal, the distribution was left-skewed, with the majority of words falling in the Low surprisal bin. There was a significant main effect of bin, F(2, 146) = 1174.55, *p* <.001, and a significant interaction between bin and group, F(6, 146) = 17.38, *p* <.001. Again, the LP group differed significantly from the other three groups across all three bins (all *ps* < 0.05).

### 3.2. Frequency trajectories

Figure 2A shows the changes in word frequency for the first 15 words in the word list, relative to the first word, for all four groups. As a baseline, we conducted a linear mixed-effect models analysis with serial position and group as fixed effects and word frequency difference as the dependent variable. Adding the interaction between serial position and group resulted in a significant improvement in model fit, *_χ_*^!^(3) = 17.96, *p* < 0.001. As an effect can be considered significant at the *p* < .05 level if the t value is well above 2 (Baayen, 2008), relative to the HP group, the LP group did not differ significantly in their frequency-by-position slope, *β* = -0.01, *t* = -0.90, whereas both the LC and HC groups showed significantly steeper frequency-by-position slopes than the HP group, *β* = -0.04, *t* = -3.73, and *β* = -0.03, *t* = -3.17, respectively.

**Figure. 2.**
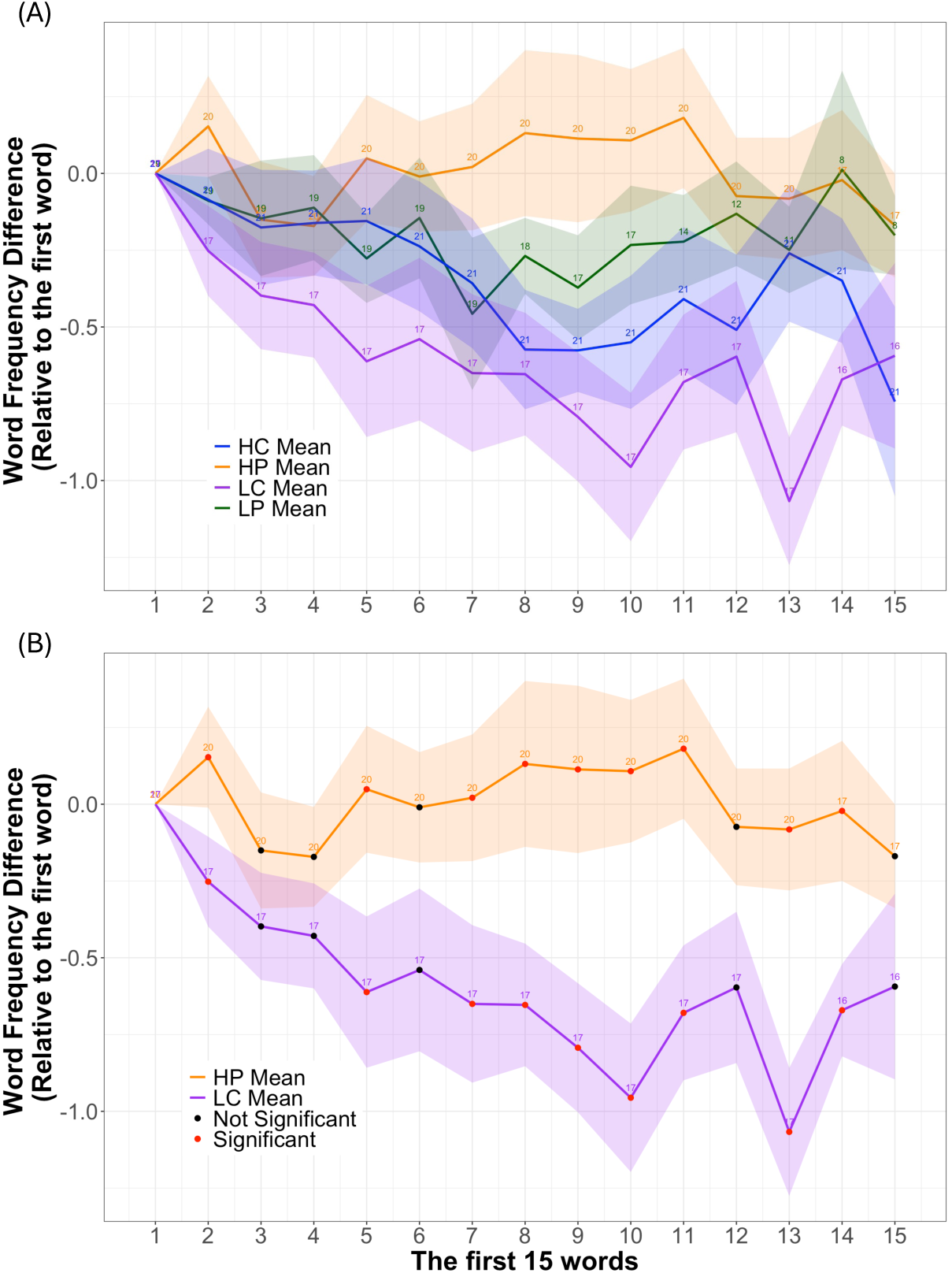
The changes of word frequency for the first 15 words in the word list for all four groups (A); and the LC and HP groups (B) with divergence analyses. The corresponding confidence intervals and sample sizes at each position are also displayed. HC: high-proficiency control; LC: low-proficiency control; HP: high-proficiency patient; LP: low-proficiency patient.

For the LC and HP group specifically, we further examined at which serial positions their frequency trajectories began to diverge. We conducted t-tests at each serial position. The LC group tended to begin the word list with high-frequency words, followed by a subsequent decline in frequency. In contrast, the HP group tended to produce words within a similar frequency band throughout the list. As shown in Figure 2B, the largest differences between the LC and HP groups were observed between fifth and eleven words. Since the number of participants contributing data across serial positions (as fewer participants produced longer lists), the corresponding confidence intervals and sample sizes at each position are displayed in Figure 2B.

### 3.3. Semantic clustering results

In the next analysis we explored one hypothesis for how the HP group was able to maintain production of equivalently frequent words across the sequence, when this metric dropped off precipitously in the other groups. If participants produce the easiest (i.e., most frequent) of the possible category items on each step, frequency would inescapably reduce across the list. One way to avoid this outcome would be for participants to switch across categories much more quickly than is typical. To test this hypothesis, we first used data-driven clustering of responses and then calculated the number of shifts that participants made across these clusters.

Figure 3 illustrates the centroid distance between each pair of the eight semantic clusters to assess their relative similarity. Lower values indicate greater semantic overlap. Overall, distances ranged from 0.11 to 0.29, reflecting relatively close semantic relationships among most clusters, consistent with all items belonging to the broader animal categories.

**Figure 3.**
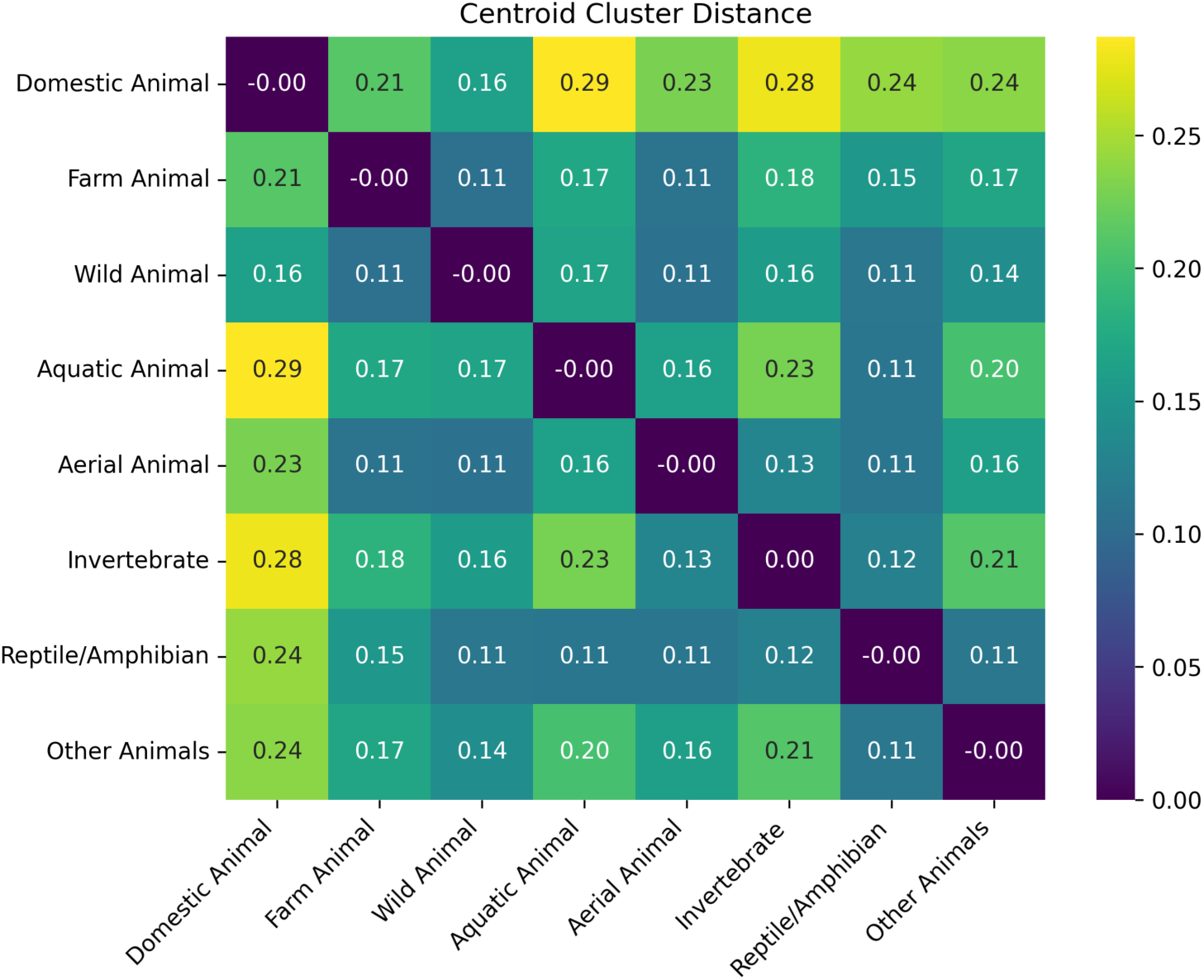
The centroid distance between each pair of the eight semantic clusters. Furthermore, the average number of movements between clusters and the average sequential cluster distance for the HP and LC groups are reported in Figure 4A and 4B, respectively. The HP group showed significantly more between-cluster movements than the LC group, *t(35)* = 2.13, *p* = 0.040. The average sequential cluster distance was also numerically higher for the HP group compared to the LC group, although the difference was only approached significant, *t(35)* = 1.77, *p* = 0.085.

**Figure 4.**
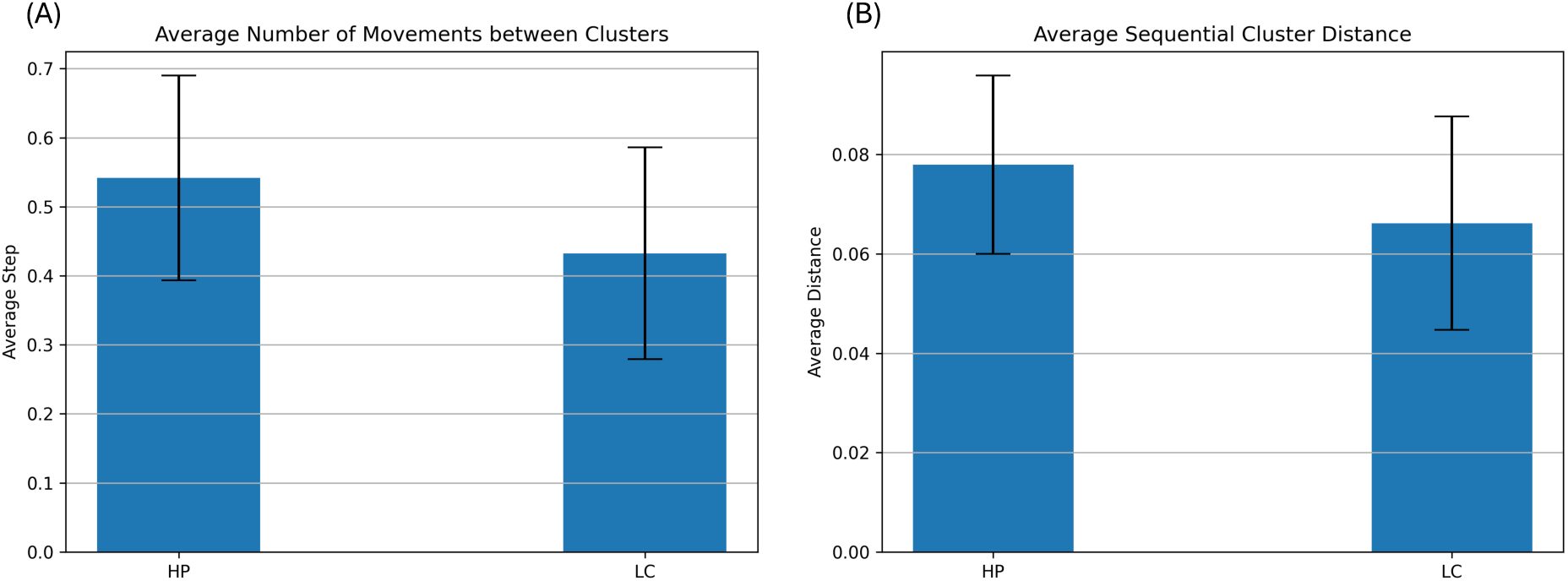
(A) The average number of movements between clusters, and (B) the average sequential cluster distance for HP and LC groups. HP: high-proficiency patient; LC: low-proficiency control.

Domestic Animal showed the greatest overall distance from other clusters, particularly Aquatic Animal (0.29) and Invertebrate (0.28), whereas Wild Animal showed the smallest distances to most other clusters, ranging from 0.11 to 0.17. This result suggests that Domestic Animal occupies a more semantically distinct position, while Wild Animal occupy a more central position relative to other categories.

### 3.5. Logistic regression analyses

We conducted two two-stage logistic regression analyses with group label as the dependent variable, one including all four groups, and the other focusing only on the HP and LC groups. In each analysis, Model 1 included only the number of unique words as a baseline predictor, while Model 2 additionally included frequency trajectory features (i.e., slope, mean and standard deviation of frequency changes across word lists) as predictor variables. The results are reported in Table 3. For the four-group analysis, the number of unique words was a significant predictor; among the frequency trajectory features, mean was a significant predictor, while slope and standard deviation were not. In contrast, for the LC-HP analysis, the number of unique words was not a significant predictor, as expected, whereas mean and standard deviation emerged as significant predictors. Critically, for both the four-group and LC-HP analyses, Model 2 could significantly improve the model fit from Model 1, *χ*^2^(3) = 19.01, *p* < .001, and *χ*^2^(3) = 18.55, *p* < .001, respectively. This pattern confirms that frequency trajectory features can explain variance beyond and above word count, particularly when groups are matched on word production proficiency.

**Table 3.** The two-stage logistic regression analyses.

|  | Four Groups |  | LC and HP Groups |  |
| --- | --- | --- | --- | --- |
|  | Model 1 | Model 2 | Model 1 | Model 2 |
|  | *Beta* | *Beta* | *Beta* | *Beta* |
| Unique word | -2.57*** | -3.74*** | -0.26 | -0.99 |
| Slope | - | 13.37 | - | 16.15 |
| Mean | - | 2.56* | - | 2.75* |
| Standard deviation | - | -6.03 | - | -8.11* |
| **Model Performance** |  |  |  |  |
| AIC | 67.04 | 54.03 | 54.48 | 41.94 |
| Accuracy | 81.8% | 87.0% | 62.16% | 81.08% |
| Kappa | 0.64 | 0.74 | 0.21 | 0.62 |
| Model Difference | < .001*** |  | < .001*** |  |

Note: \*\*\* $p < .001$ ; \*\* $p < .01$ ; \* $p < .05$

To further investigate the classification performance of the regression models, we plotted receiver operating characteristic (ROC) curves for both analyses, as shown in Figure 5. The area under the curve (AUC) values indicated that including frequency trajectory information substantially improved the model’s classification performance in both analyses (four-group analysis: AUC = 0.906 for Model 1 vs. 0.947 for Model 2; LC–HP analysis: AUC = 0.706 for Model 1 vs. 0.876 for Model 2).

**Figure 5.**
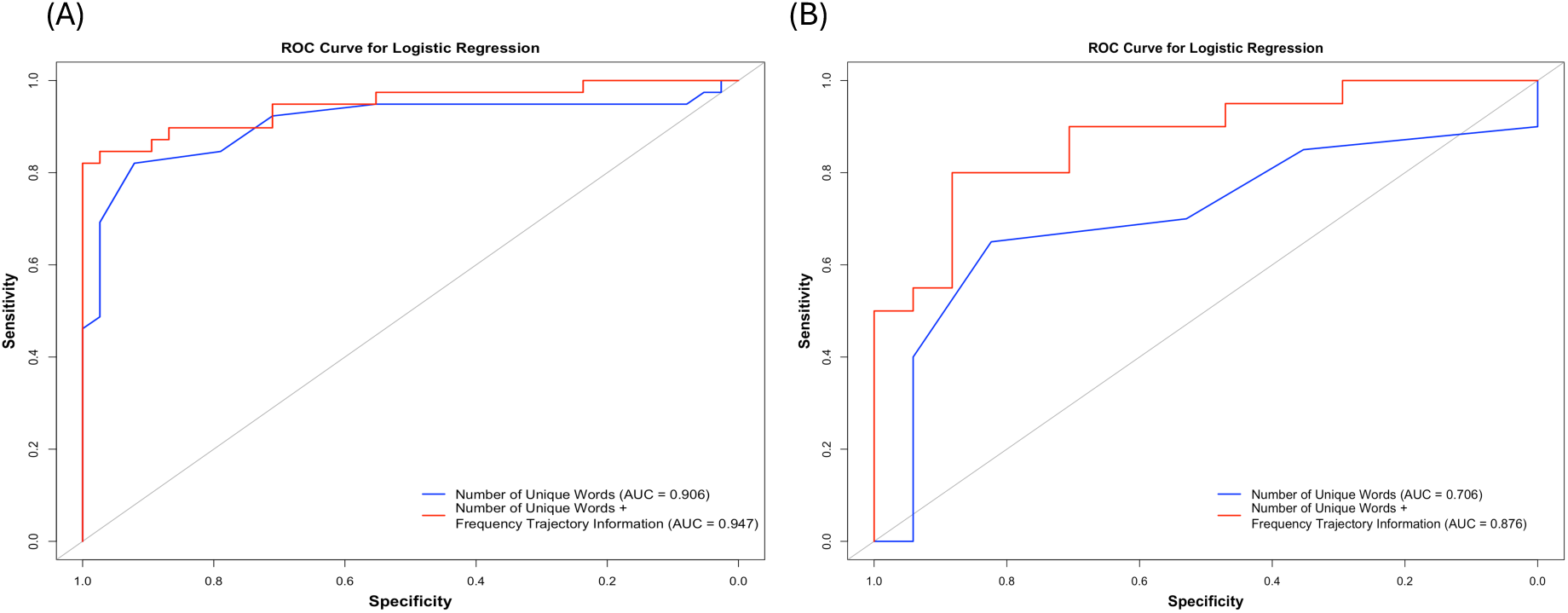
The receiver operating characteristic (ROC) curves for both logistic regression analyses on all the four groups (HC, HP, LC and LP) (A); and only on the HP and LC groups. HC: high-proficiency control; LC: low-proficiency control; HP: high-proficiency patient; LP: low-proficiency patient.

Finally, we evaluated the generalisability of Model 2 from the four-group analysis using an independent out-of-sample dataset. The model achieved an accuracy of 85.19%, a sensitivity of 90.48%, and a specificity of 66.67%. The substantial difference between sensitivity and specificity likely reflects the class imbalance between health controls and patients in this out-of-sample set.

## 4. Discussion

Numerous studies have demonstrated that word count produced during VF tasks is a key indicator of cognitive impairment and neurodegenerative diseases (McDonnell et al., 2020; Mueller et al., 2015). Although simple to administer and score, word count alone struggles to differentiate high proficient patients, who can produce a similar number of words as healthy individuals. Our study addressed this challenge by investigating the item-level and sequential properties of words produced by healthy individuals and the patient groups. The results demonstrated that many item-level word properties could distinguish the LP group from other groups, consistent with previous work (Forbes-McKay et al., 2005; Henderson et al. 2023; Wakefield et al., 2018), however these did not reliably differentiate the LC and HP groups. Instead, the sequential properties of words, specifically, frequency trajectories, were able to detect subtle differences between the LC and HP groups.

As shown in Figure 1, the LP group predominantly retrieved the most common, contextually familiar, and predictable animals, with little word-frequency diversity in their lexical search. In contrast, the HC, LC, and HP groups showed a more evenly spread distribution across word-frequency bins, reflecting a broader range of words retrieved. Despite this overall difference driven by the LP group, the HC, LC, and HP groups exhibited broadly similar distributional profiles to one another, suggesting that psycholinguistic properties of the words alone are insufficient to differentiate the LC and HP groups. This limited discriminability is consistent with findings from Henderson et al. (2023), who examined whether total word count and psycholinguistic word properties produced during VF tasks could reliably differentiate healthy controls from patients across six neurodegenerative conditions. While total word count successfully distinguished controls from all patient groups, neither this measure nor the psycholinguistic properties of the words produced could differentiate between patient groups, with the partial exception of SD/svPPA. Taken together, these findings suggest that psycholinguistic properties of the words produced during VF tasks may only be sensitive to more serve or diagnostically deviant patient profiles and have limited utility for detecting subtler impairments such as those seen in high-proficiency cognitive impaired patients.

The current study suggests that a more profitable approach may be to consider the sequential patterns in the patients’ response rather than analyse them independently of each other. Specifically, we found that while the psycholinguistic properties of words produced during VF tasks were insufficient to differentiate the LC and HP groups, frequency trajectories provided critical discrimination information. As illustrated in Figure 2, the LC and HP groups could be clearly separated by their frequency trajectories, where the HP group tended to produce words within a relatively narrow frequency ranges throughout the list, whereas the LC group tended to begin with high-frequency words and show a systematic decline in frequency across serial positions. These differences were most pronounced between the fifth and eleven words. Semantic cluster analyses further revealed that the HP group made significantly more between-cluster transitions than the LC group, reflecting qualitatively different patterns of lexical retrieval. Specifically, the HP patients appear to retrieve words by ranging broadly across different semantic clusters while maintaining a relatively stable frequency band, rather than exhausting high-frequency words within one cluster before transitioning to another. In contrast, LC controls (and the other groups) appear to begin with the most common exemplars within a given semantic cluster, gradually retrieving lower-frequency items before switching to a new cluster. This pattern is broadly consistent with the clustering and switching framework (Troyer et al., 1997; Weakley et al., 2014), in which efficient retrieval involves systematic within-cluster search followed by strategic between-cluster transitions. The present findings suggest that high-proficiency patients may adopt a different retrieval strategy, that prioritises lexical familiarity over systematic semantic search, reflecting subtle differences in the organisation or accessibility of semantic memory and access to the names of the concepts.

The logistic regression results further confirmed these findings. The number of unique words was a strong discriminator in the four-group analysis (Model 1: AUC =0.906), however, its discriminability dropped substantially when the analysis was restricted to the LC and HP groups (Model 1 AUC = 0.706). Critically, the inclusion of frequency trajectory features improved the AUC by approximate 4.1% for the four-group analysis (Model 2 AUC = 0.947) and by 17% for the LC-HP analysis (Model 2 AUC = 0.876), demonstrating that frequency trajectory information provides meaningful discriminative value above and beyond unique word count, particularly in cases where word production proficiency is comparable across groups. Furthermore, the four-group model achieved an accuracy of 85.2%, a sensitivity of 90.5%, and a specificity of 66.7% on the out-of-sample test, suggesting adequate generalisation to an independent sample. Collectively, these results indicate that incorporating sequential word frequency information into the analysis of VF performance holds promise as a clinically applicable approach for detecting patients with cognitive impairment, even in patients whose overall word production appears intact.

Several limitations should be considered. First, our sample sizes were relatively small, particularly in the out-of-sample dataset, which was also class-imbalanced. Future studies should recruit larger and more balanced samples to improve the robustness and generalisability of the findings. Second, our study focused on the psycholinguistic properties of Mandarin Chinese words. As such word properties may vary widely across language systems, it would be valuable to examine whether frequency trajectory patterns observed here generalise cross-linguistically, particularly to languages with different orthographic and phonological structures. Third, our participant data were cross-sessional, and due to limited data, we could not examine the longitudinal changes in participants’ performance in VF tasks. Future work could collect and leverage longitudinal data to track whether low-proficiency controls show a higher rate of conversion to cognitive impairment over time. This would further validate the clinical utility of frequency trajectory features as an early indicator of cognitive decline, potentially identifying at-risk individuals even before their word production proficiency declines to patient levels.

## 5. Conclusion

This study investigated the item-level and sequential properties of words produced by healthy individuals and the patients with cognitive impairment during a VF task. The results suggest that psycholinguistic properties of words independently of each other could only distinguish the LP group from other groups, while frequency trajectories could detect subtle differences between patients and controls with comparable levels of word production proficiency, reflecting differences in the sequential organisation of semantic access. Critically, these differences were most pronounced at serial positions five to eleven and adequately generalised to an out-of-sample dataset, suggesting that sequential word properties may offer a sensitive and efficient marker for detecting even when overall fluency performance appears intact.

## Acknowledgement

We thank our patients and their families for supporting this work.

## Data availability statement

The authors confirm that the data supporting the findings of this study are available within the article. Additional data are available from the corresponding author upon reasonable request.

## Ethics statement

The studies involving humans were approved by Institutional Review Board of Cardinal Tien Hospital. The studies were conducted in accordance with the local legislation and institutional requirements. Written informed consent for participation in this study was provided by the participants’ legal guardians or next of kin.

